# Population age as a key factor in the COVID-19 pandemic dynamics

**DOI:** 10.1101/2023.11.30.23299229

**Authors:** Igor Nesteruk, Matt Keeling

**Affiliations:** Institute of Hydromechanics, National Academy of Sciences of Ukraine, Kyiv, Ukraine, SBIDER (Systems Biology & Infectious Disease Epidemiology Research) Centre at, University of Warwick, UK; School of Life Sciences & Mathematics Institute, University of Warwick, UK

**Keywords:** COVID-19 pandemic dynamics, Zero-COVID strategy, numbers of cases and deaths per capita, case fatality risk, mathematical modeling of infection diseases, statistical methods

## Abstract

The population and governments of many countries are losing interest in the SARS-CoV-2 infection, the number of tests and the number of new cases detected is sharply decreasing. To compare the accumulated numbers *CC* of cases and deaths *DC* per million and to answer the question why the less vaccinated Africa has accumulated 36 times lower *CC* values and 15 times lower *DC* values than Europe, simple statistical analysis have been performed. *CC* and *DC* values demonstrated rather strong correlation with the median age of populations. One-year increment in the median year yields 12-18 thousand increase in *CC* values and 52-83 increase in *DC* values. Zero-COVID countries succeed to have much lower numbers of deaths per capita and case fatality ratios *DC/CC*.

---

The fourth year of the COVID-19 pandemic [1, 2] is characterized by a high number of re-infections [3-5] associated with the circulation of new strains of SARS-CoV-2 [6-8] and an increase in the case fatality risk *CFR* (in 2023 the global *CFR* values were almost twice higher than in 2022 [9]). Nevertheless, the population and governments of many countries are losing interest in the infection, the number of tests and the number of new cases detected is sharply decreasing. In particular, the US, China and Japan have stopped to update their COVID-19 datasets after May 15, 2023; Turkey has reported no new cases in 2023, [1, 2]. Before the data update is finally stopped, it would be very interesting to compare the accumulated numbers *CC* of cases and deaths *DC* per capita and to answer the question why the less vaccinated Africa has accumulated 36 times lower *CC* values and 15 times lower *DC* values than Europe (see [1, 2], lines 74 and 75 in Table S1). This information, as well as *CFR* values, would make it possible to evaluate the methods of combating the pandemic, and to assess trends in pandemic dynamics, in particular, its relationship with the age of the populations.

The severity of SARS-CoV-2 infection (in particular, the need for oxygen supplementation) increases for older patients [10]. Younger populations have less clinical cases per capita [11]. Almost half of the infected children less than 15 years old can be asymptomatic [12]. Together with many asymptomatic adult patients [13 - 17] they can make the main part of pandemic dynamics invisible [18]. Paper [11] (published in 2020) predicted that “Without effective control measures, regions with relatively older populations could see disproportionally more cases of COVID-19, particularly in the later stages of an unmitigated epidemic”. In this study we will verify this hypothesis and analyze the results of Zero-COVID strategy which “involves using public health measures such as contact tracing, mass testing, border quarantine, lockdowns, and mitigation software in order to stop community transmission of COVID-19 as soon as it is detected”, [19]. The goal of Zero-COVID strategy is to “get the area back to zero new infections and resume normal economic and social activities” [19]. It was applied in different countries mostly in Asia and Oceania in 2020, 2021 and early 2022.

We will use the accumulated numbers of laboratory-confirmed COVID-19 cases *CC*_*i*_ and deaths *DC*_*i*_ per million (*i* =1, 2,.., 79) for some countries, regions, continents, and the world listed in COVID-19 Data Repository by the Center for Systems Science and Engineering (CSSE) at Johns Hopkins University (JHU), [2] (version of file updated on September 28, 2023, see supplementary Table S1). The have chosen the countries with the population larger than 20 millions (according [20]; all figures correspond to October 11, 2023) making exception for Ukraine, Syria, Afghanistan, Yemen, and Russia to avoid influence of military operations on the COVID-19 statistics. There is no data from North Korea. Since Chinese statistics shows some contradictions (see, e.g., [21] or compare JHU files updated on September 28 and March 9, 2023), we have used only figures for Taiwan and Hong Kong. In particular, in Table S1 we show corresponding *CC*_*i*_ and *DC*_*i*_ values from the March-9-version of JHU file (not available on September 28). The *CC*_*i*_ and *DC*_*i*_ values for Taiwan and Hong Kong for 2023 we have calculated with the use of [22-25].

We have added the figures corresponding to some countries with population less than 20 millions representing different regions and information about 10 Zero-COVID countries and regions (see lines 63-72 in Table S1). It must be noted that aggregated values of *CC*_*i*_ and *DC*_*i*_ corresponding to EU, continents and the world (see lines 73-79 in Table S1) contain the information about all the countries (in particular, Ukrainian and Chinese datasets). We have listed also the information about the median ages *A*_*i*_ from [20, 26] (see Table S1).

The case fatality risks can be calculated by dividing corresponding *DC*_*i*_ values by *CC*_*i*_ according to the formula *CFR*_*i*_ =(*DC*_*i*_/*CC*_*i*_)*100%. The results of calculations are also listed in Table S1. We will use the linear regression and Fisher test [27] to investigate the links between age *A* and *CC, DC* and *CFR* values (see Supplementary). The numbers of observations *n* will correspond to the countries listed in lines 1-62 of Table S1 (*n*=62), Zero-COVID countries (lines 63-72, *n*=10), and total dataset including continents and the world (*n*=79).

The results of calculations are listed in Table 1 and illustrated in Fig. 1 with the use of small markers and thin lines for values accumulated before December 31, 2022 and large markers and bold lines for values accumulated before September 10, 2023. Since the differences in best fitting lines for *CRF* calculated with the use of 2022 and 2023 datasets are very small (compare values of parameters *a* and *b* listed in Table 1, lines 13-18), we show only bold lines corresponding to September 10, 2023. Blue color corresponds to the accumulated numbers of cases per ten persons (*CC*/100,000), black color - to the accumulated numbers of deaths per thousand (*DC*/1000) and red one - to the *CFR* values in %. “Circles” show the data for Zero-COVID countries, “squares” – aggregated characteristics for EU, continents and the world.

**Table 1.** Optimal values of parameters in eq. (S1), correlation coefficients and the results of Fisher test applications.

| No. | Dataset | Number of observations<br>$n$ | Correlation coefficient<br>$R$ | Optimal values of parameter $a$<br>in eq. (S1) | Optimal values of parameter $b$<br>in eq. (S1) | Experimental value of the Fisher function $F$, eq. (S2), $m=2$ | Critical value of Fisher function $F_c(I, n-2)$ for the confidence level 0.05, [28] | $F/F_c$ |
| --- | --- | --- | --- | --- | --- | --- | --- | --- |
| **Correlations for numbers of COVID-19 cases per million accumulated as of December 31, 2022, $CC^{(1)}$** |  |  |  |  |  |  |  |  |
| 1 | Countries S1, 1-62 | 62 | 0.7264 | -230313.1 | 12067.305 | 67.04 | 4.0 | 16.8 |
| 2 | Zero-COVID | 10 | 0.7536 | -245204.0 | 14918.425 | 10.51 | 5.32 | 2.0 |
| 3 | Total | 79 | 0.7675 | -258318.2 | 13385.145 | 110.40 | 3.98 | 27.8 |
| **Correlations for numbers of COVID-19 cases per million accumulated as of September 10, 2023, $CC^{(2)}$** |  |  |  |  |  |  |  |  |
| 4 | Countries S1, 1-62 | 62 | 0.7308 | -238708.4 | 12464.814 | 68.78 | 4.0 | 17.2 |
| 5 | Zero-COVID | 10 | 0.7348 | -310865.6 | 17640.497 | 9.39 | 5.32 | 1.8 |
| 6 | Total | 79 | 0.7653 | -277002.4 | 14248.047 | 108.83 | 3.98 | 27.4 |
| **Correlations for numbers deaths per million accumulated as of December 31, 2022, $DC^{(1)}$** |  |  |  |  |  |  |  |  |
| 7 | Countries S1,1-62 | 62 | 0.5878 | -1222.941 | 80.0912 | 31.667 | 4.0 | 7.9 |
| 8 | Zero-COVID | 10 | 0.6765 | -1277.277 | 51.7604 | 6.75 | 5.32 | 1.3 |
| 9 | Total | 79 | 0.5361 | -1009.386 | 69.4481 | 31.06 | 3.98 | 7.8 |
| **Correlations for numbers deaths per million accumulated as of September 10, 2023, $DC^{(2)}$** |  |  |  |  |  |  |  |  |
| 10 | Countries S1, 1-62 | 62 | 0.5983 | -1292.541 | 83.4639 | 33.45 | 4.0 | 8.4 |
| 11 | Zero-COVID | 10 | 0.7058 | -1448.283 | 58.9263 | 7.94 | 5.32 | 1.5 |
| 12 | Total | 79 | 0.5550 | -1095.528 | 73.5575 | 34.28 | 3.98 | 8.6 |
| **Correlations for the case fatality risks as of December 31, 2022, $CFR^{(1)}$** |  |  |  |  |  |  |  |  |
| 13 | Countries S1, 1-62 | 62 | -0.3885 | 3.21772 | -0.05371 | 10.67 | 4.0 | 2.7 |
| 14 | Zero-COVID | 10 | 0.4260 | -0.16976 | 0.01022 | 1.77 | 5.32 | 0.3 |
| 15 | Total | 79 | -0.4420 | 3.22287 | -0.05871 | 18.70 | 3.98 | 4.7 |
| **Correlations for the case fatality risks as of September 10, 2023, $CFR^{(2)}$** |  |  |  |  |  |  |  |  |
| 16 | Countries S1, 1-62 | 62 | -0.3839 | 3.18254 | -0.05278 | 10.37 | 4.0 | 2.6 |
| 17 | Zero-COVID | 10 | 0.4430 | -0.18372 | 0.01085 | 1.95 | 5.32 | 0.4 |
| 18 | Total | 79 | -0.4374 | 3.18636 | -0.05772 | 18.22 | 3.98 | 4.6 |
| 19 | Countries S1, 1-62, $A_i < 30$ | 34 | 0.0207 | 2.00123 | 0.005783 | 0.014 | 4.16 | 0.0033 |

**Fig. 1.**
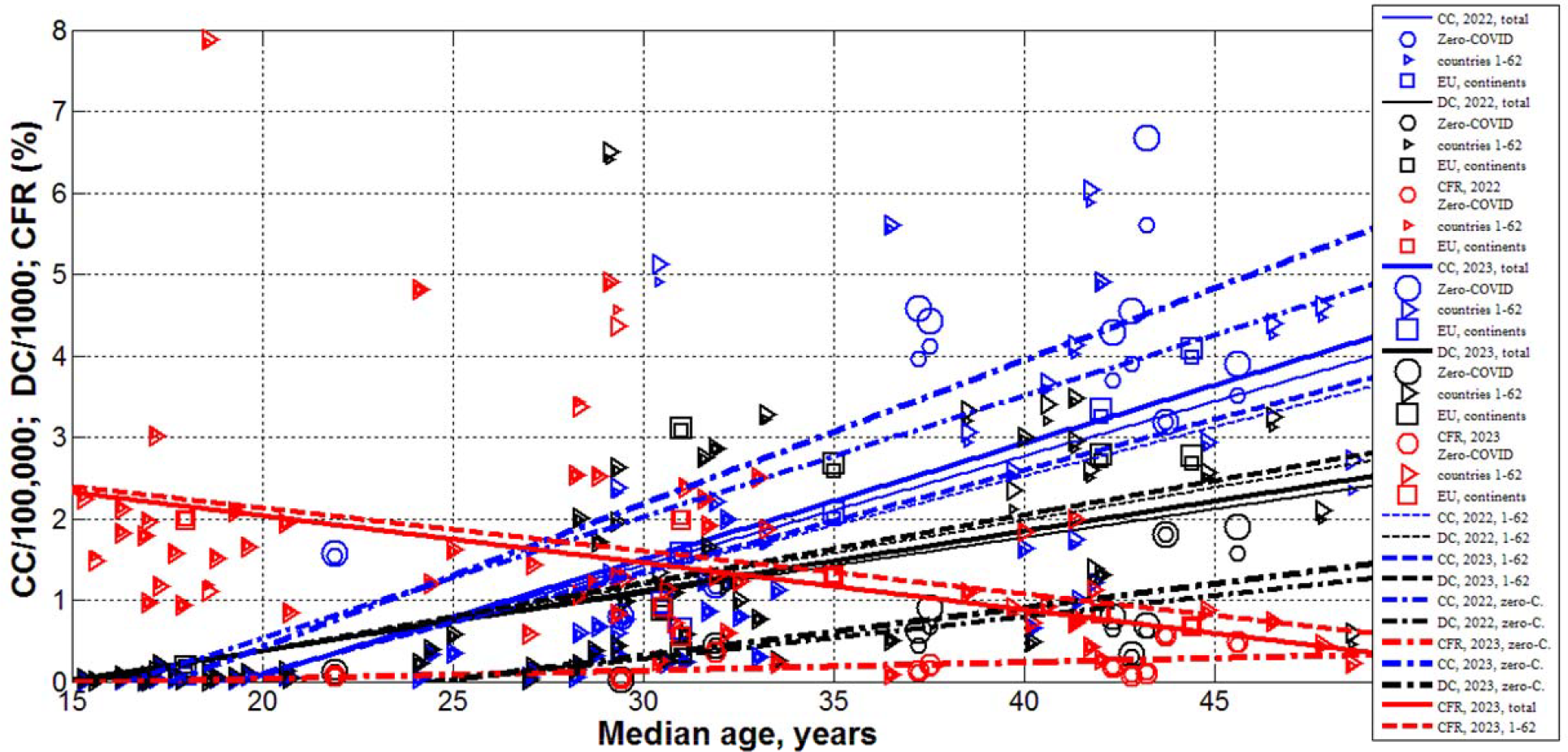
Accumulated numbers of COVID-19 cases per ten persons (*CC*/100,000; blue), accumulated numbers of deaths per thousand (*DC*/1000; black), case fatality risks (*CFR*, %, red), and best fitting lines versus median age in years. Bolder lines and larger markers correspond to September 10, 2023. Solid lines were calculated with the use of total dataset listed in Table S1, dashed lines – only to countries listed in lines 1-62, dashed-dotted lines – to Zero-COVID countries listed in lines 63-72.

The accumulated numbers of cases per capita demonstrate rather strong correlation with the median age. High values of the correlation coefficient in lines 1-6 of Table 1 show that more than 52% of variations in *CC* are connected with the age. The values of the parameter *b* (eq. S1) listed in these lines demonstrate that one-year increment in the median year yields 12-18 thousand increase in *CC* values. Probably due to contact tracing and mass testing, Zero-COVID countries have accumulated more cases per capita (compare dashed-dotted blue lines with solid and dashed blue lines).

The accumulated numbers of deaths per capita demonstrate weaker correlation with the median age. The values of the correlation coefficient in lines 7-12 of Table 1 show that only 29-50% of variations in *DC* are related to the age. The values of the parameter *b* demonstrate that one-year increment in the median year yields 52-83 increase in *DC* values. Zero-COVID countries succeed to have much lower numbers of deaths per capita (compare dashed-dotted black lines with solid and dashed black lines).

The Fisher test supports the growth of *CC* and *DC* values with the increase of median age. Even for small number of Zero-COVID countries (*n*=10), the correlations are valid at the significance level 0.05 (see last values in lines 1-12 of Table 1). For other countries the correlations were supported at the much higher significance levels (e.g., 0.001). Thus, we really see disproportionally more COVID-19 cases and deaths in regions with older populations. Even in Zero-COVID countries, where special and sometimes very severe lockdowns and control measures were applied, we see the same correlation. Nevertheless, these countries succeed to reduce the case fatality risks significantly (compare red best fitting lines).

Since in 2020 younger populations had less clinical cases per capita [11], it was expected to obtain lower *CFR* values as well. Nevertheless, rows 13, 15, 16, 18 in Table 1 and red solid and dashed lines indicate the opposite trend (for Zero-COVID countries no correlations were revealed, see rows 14 and 17). Lower values of *r*^*2*^ (in comparison with correlations for *CC* and *DC*) allow concluding that the influence of age on case fatality risk is rather weak. For example, no correlation was revealed for the countries with the median age less than 30 years (see line 19 in Table 1). The probability that a person tested positive will die (i.e., *CFR*) depends not only on the individual state of health and immunity (related to age and vaccinations), but also to a large extent on the speed and quality of medical care, which are better in countries with higher incomes (and older populations). As of August 1, 2022, the *DC* and *CFR* values decreased for richer European countries [29]. Probably, the better medical care and higher vaccination level in richer countries helps to reduce *CFR* despite older populations.

## Clarification point

No humans or human data was used during this study

## Data availability

All data generated or analyzed during this study are included in this text.

## Acknowledgements

The study was supported by the Solidarity Satellite Programme of Isaac Newton Institute for Mathematical Sciences, Cambridge, UK. The authors are grateful to Professor Robin Thompson and Oleksii Rodionov for their support and providing very useful information.

## Supplementary

The linear regression is used to calculate the regression coefficients *r* and the coefficients *a* and *b* of corresponding best fitting straight lines, [27]:

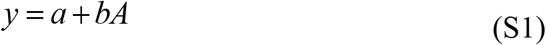

where explanatory variable is median age *A* and dependent variables *y* are *CC, DC* and *CFR*.

The F-test for the null hypothesis that says that the proposed linear relationship (S1) fits the data sets. The experimental values of the Fisher function can be calculated with the use of the formula:

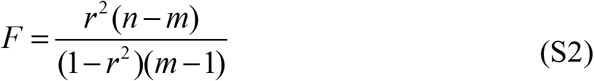

where *n* is the number of observations (number of countries and regions taken for statistical analysis); *m*=2 is the number of parameters in the regression equation, [27]. The corresponding experimental values *F* have to be compared with the critical values *F*_*C*_ (*k*_1_, *k*_2_) of the Fisher function at a desired significance or confidence level (*k*_1_ = *m* −1, *k*_2_ = *n* − *m*, see, e.g., [28]). If *F* / *F*_*C*_ (*k*_1_, *k*_2_) <1, the null hypothesis is not supported by the results of observations. The highest values of reliable correlations. *F* / *F*_*C*_ (*k*_1_, *k*_2_) correspond to the most reliable correlations.

**Table S1.**
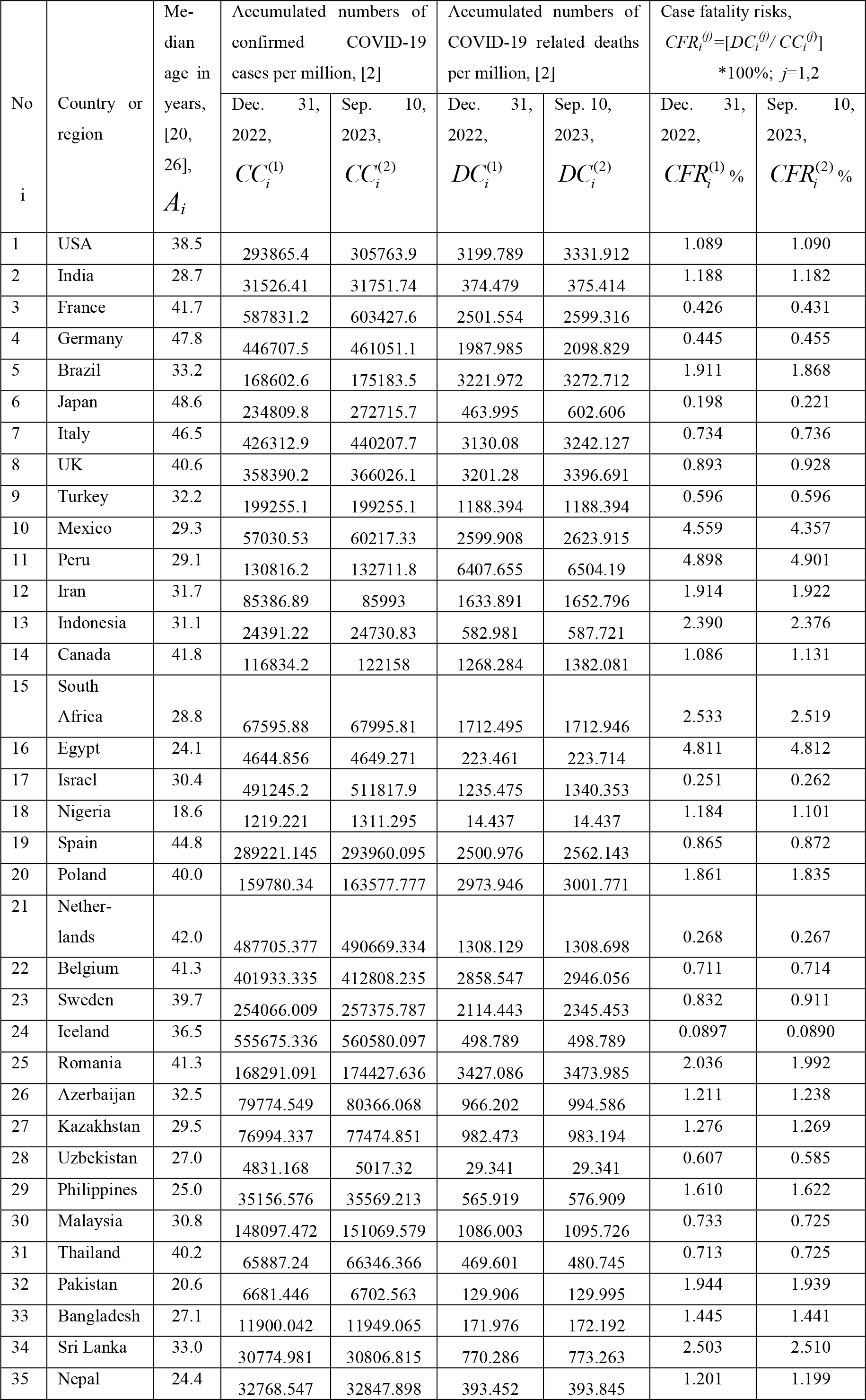
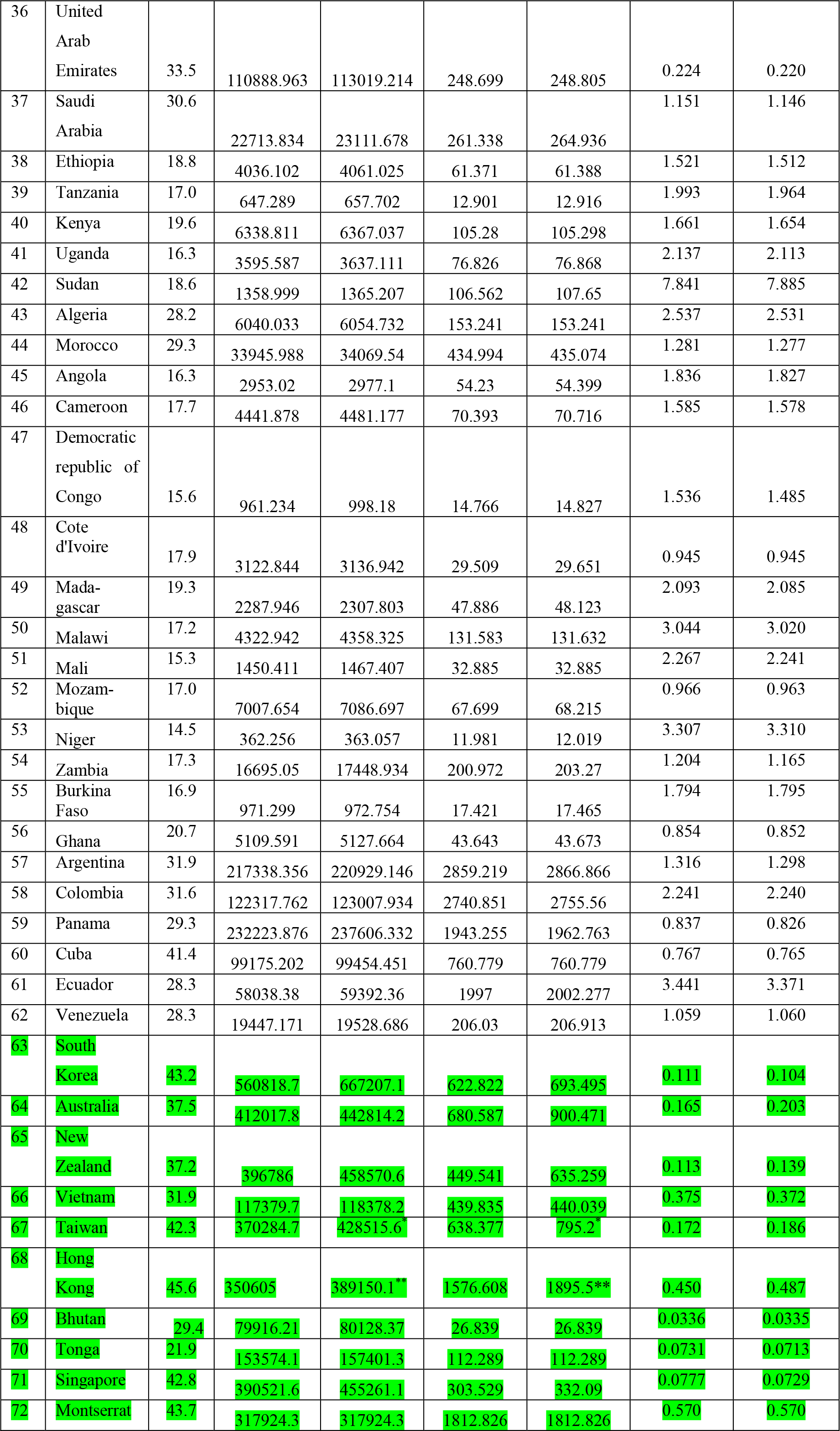
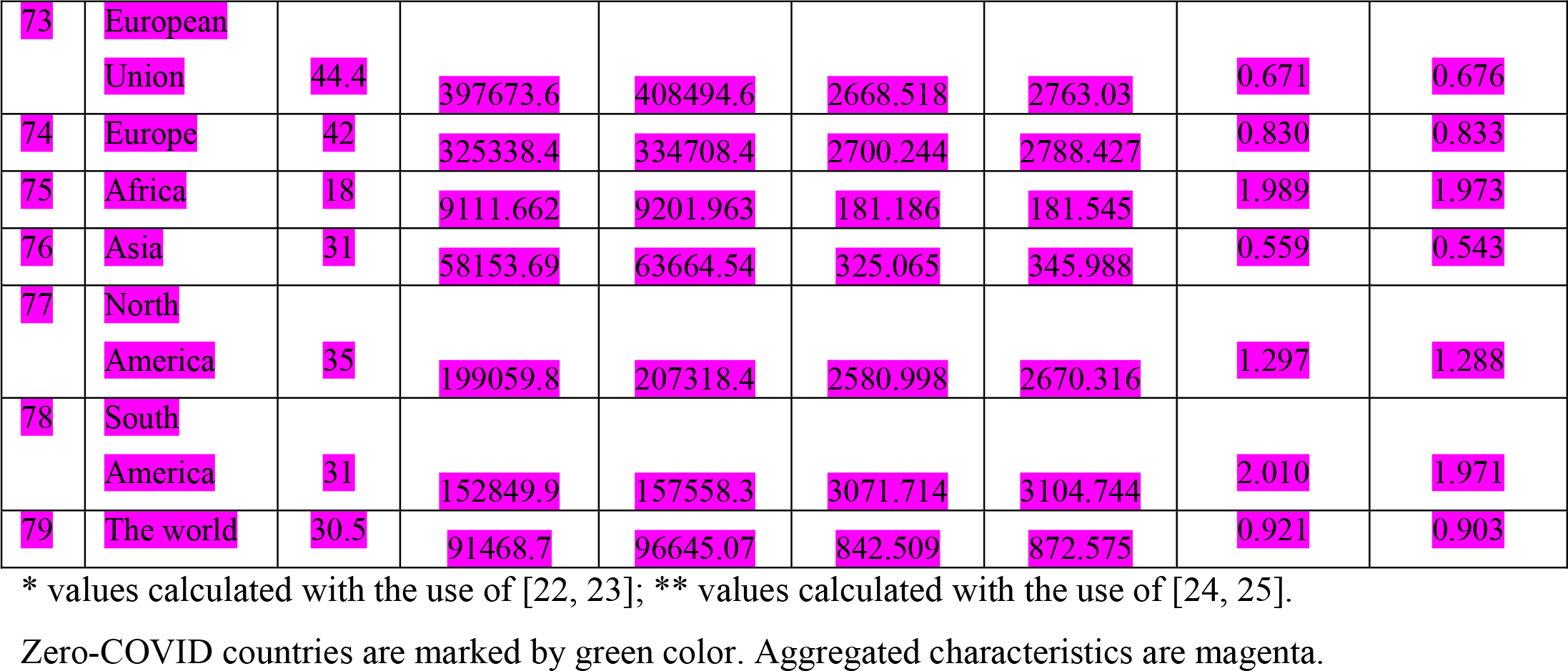
Median age, accumulated numbers the COVID-19 cases and deaths per capita, case fatality risks.

| No<br>i | Country or<br>region | Me-<br>dian<br>age in<br>years,<br>[20,<br>26],<br>$A_i$ | Accumulated numbers of<br>confirmed COVID-19<br>cases per million, [2] | | Accumulated numbers of<br>COVID-19 related deaths<br>per million, [2] | | Case fatality risks,<br>$CFR_i^{(j)}=[DC_i^{(j)}/CC_i^{(j)}]$<br>*100%; $j=1,2$ | |
| --- | --- | --- | --- | --- | --- | --- | --- | --- |
| | | | Dec. 31,<br>2022,<br>$CC_i^{(1)}$ | Sep. 10,<br>2023,<br>$CC_i^{(2)}$ | Dec. 31,<br>2022,<br>$DC_i^{(1)}$ | Sep. 10,<br>2023,<br>$DC_i^{(2)}$ | Dec. 31,<br>2022,<br>$CFR_i^{(1)}\%$ | Sep. 10,<br>2023,<br>$CFR_i^{(2)}\%$ |
| 1 | USA | 38.5 | 293865.4 | 305763.9 | 3199.789 | 3331.912 | 1.089 | 1.090 |
| 2 | India | 28.7 | 31526.41 | 31751.74 | 374.479 | 375.414 | 1.188 | 1.182 |
| 3 | France | 41.7 | 587831.2 | 603427.6 | 2501.554 | 2599.316 | 0.426 | 0.431 |
| 4 | Germany | 47.8 | 446707.5 | 461051.1 | 1987.985 | 2098.829 | 0.445 | 0.455 |
| 5 | Brazil | 33.2 | 168602.6 | 175183.5 | 3221.972 | 3272.712 | 1.911 | 1.868 |
| 6 | Japan | 48.6 | 234809.8 | 272715.7 | 463.995 | 602.606 | 0.198 | 0.221 |
| 7 | Italy | 46.5 | 426312.9 | 440207.7 | 3130.08 | 3242.127 | 0.734 | 0.736 |
| 8 | UK | 40.6 | 358390.2 | 366026.1 | 3201.28 | 3396.691 | 0.893 | 0.928 |
| 9 | Turkey | 32.2 | 199255.1 | 199255.1 | 1188.394 | 1188.394 | 0.596 | 0.596 |
| 10 | Mexico | 29.3 | 57030.53 | 60217.33 | 2599.908 | 2623.915 | 4.559 | 4.357 |
| 11 | Peru | 29.1 | 130816.2 | 132711.8 | 6407.655 | 6504.19 | 4.898 | 4.901 |
| 12 | Iran | 31.7 | 85386.89 | 85993 | 1633.891 | 1652.796 | 1.914 | 1.922 |
| 13 | Indonesia | 31.1 | 24391.22 | 24730.83 | 582.981 | 587.721 | 2.390 | 2.376 |
| 14 | Canada | 41.8 | 116834.2 | 122158 | 1268.284 | 1382.081 | 1.086 | 1.131 |
| 15 | South<br>Africa | 28.8 | 67595.88 | 67995.81 | 1712.495 | 1712.946 | 2.533 | 2.519 |
| 16 | Egypt | 24.1 | 4644.856 | 4649.271 | 223.461 | 223.714 | 4.811 | 4.812 |
| 17 | Israel | 30.4 | 491245.2 | 511817.9 | 1235.475 | 1340.353 | 0.251 | 0.262 |
| 18 | Nigeria | 18.6 | 1219.221 | 1311.295 | 14.437 | 14.437 | 1.184 | 1.101 |
| 19 | Spain | 44.8 | 289221.145 | 293960.095 | 2500.976 | 2562.143 | 0.865 | 0.872 |
| 20 | Poland | 40.0 | 159780.34 | 163577.777 | 2973.946 | 3001.771 | 1.861 | 1.835 |
| 21 | Nether-<br>lands | 42.0 | 487705.377 | 490669.334 | 1308.129 | 1308.698 | 0.268 | 0.267 |
| 22 | Belgium | 41.3 | 401933.335 | 412808.235 | 2858.547 | 2946.056 | 0.711 | 0.714 |
| 23 | Sweden | 39.7 | 254066.009 | 257375.787 | 2114.443 | 2345.453 | 0.832 | 0.911 |
| 24 | Iceland | 36.5 | 555675.336 | 560580.097 | 498.789 | 498.789 | 0.0897 | 0.0890 |
| 25 | Romania | 41.3 | 168291.091 | 174427.636 | 3427.086 | 3473.985 | 2.036 | 1.992 |
| 26 | Azerbaijan | 32.5 | 79774.549 | 80366.068 | 966.202 | 994.586 | 1.211 | 1.238 |
| 27 | Kazakhstan | 29.5 | 76994.337 | 77474.851 | 982.473 | 983.194 | 1.276 | 1.269 |
| 28 | Uzbekistan | 27.0 | 4831.168 | 5017.32 | 29.341 | 29.341 | 0.607 | 0.585 |
| 29 | Philippines | 25.0 | 35156.576 | 35569.213 | 565.919 | 576.909 | 1.610 | 1.622 |
| 30 | Malaysia | 30.8 | 148097.472 | 151069.579 | 1086.003 | 1095.726 | 0.733 | 0.725 |
| 31 | Thailand | 40.2 | 65887.24 | 66346.366 | 469.601 | 480.745 | 0.713 | 0.725 |
| 32 | Pakistan | 20.6 | 6681.446 | 6702.563 | 129.906 | 129.995 | 1.944 | 1.939 |
| 33 | Bangladesh | 27.1 | 11900.042 | 11949.065 | 171.976 | 172.192 | 1.445 | 1.441 |
| 34 | Sri Lanka | 33.0 | 30774.981 | 30806.815 | 770.286 | 773.263 | 2.503 | 2.510 |
| 35 | Nepal | 24.4 | 32768.547 | 32847.898 | 393.452 | 393.845 | 1.201 | 1.199 |
| 36 | United Arab Emirates | 33.5 | 110888.963 | 113019.214 | 248.699 | 248.805 | 0.224 | 0.220 |
| 37 | Saudi Arabia | 30.6 | 22713.834 | 23111.678 | 261.338 | 264.936 | 1.151 | 1.146 |
| 38 | Ethiopia | 18.8 | 4036.102 | 4061.025 | 61.371 | 61.388 | 1.521 | 1.512 |
| 39 | Tanzania | 17.0 | 647.289 | 657.702 | 12.901 | 12.916 | 1.993 | 1.964 |
| 40 | Kenya | 19.6 | 6338.811 | 6367.037 | 105.28 | 105.298 | 1.661 | 1.654 |
| 41 | Uganda | 16.3 | 3595.587 | 3637.111 | 76.826 | 76.868 | 2.137 | 2.113 |
| 42 | Sudan | 18.6 | 1358.999 | 1365.207 | 106.562 | 107.65 | 7.841 | 7.885 |
| 43 | Algeria | 28.2 | 6040.033 | 6054.732 | 153.241 | 153.241 | 2.537 | 2.531 |
| 44 | Morocco | 29.3 | 33945.988 | 34069.54 | 434.994 | 435.074 | 1.281 | 1.277 |
| 45 | Angola | 16.3 | 2953.02 | 2977.1 | 54.23 | 54.399 | 1.836 | 1.827 |
| 46 | Cameroon | 17.7 | 4441.878 | 4481.177 | 70.393 | 70.716 | 1.585 | 1.578 |
| 47 | Democratic republic of Congo | 15.6 | 961.234 | 998.18 | 14.766 | 14.827 | 1.536 | 1.485 |
| 48 | Cote d'Ivoire | 17.9 | 3122.844 | 3136.942 | 29.509 | 29.651 | 0.945 | 0.945 |
| 49 | Mada-gascar | 19.3 | 2287.946 | 2307.803 | 47.886 | 48.123 | 2.093 | 2.085 |
| 50 | Malawi | 17.2 | 4322.942 | 4358.325 | 131.583 | 131.632 | 3.044 | 3.020 |
| 51 | Mali | 15.3 | 1450.411 | 1467.407 | 32.885 | 32.885 | 2.267 | 2.241 |
| 52 | Mozam-bique | 17.0 | 7007.654 | 7086.697 | 67.699 | 68.215 | 0.966 | 0.963 |
| 53 | Niger | 14.5 | 362.256 | 363.057 | 11.981 | 12.019 | 3.307 | 3.310 |
| 54 | Zambia | 17.3 | 16695.05 | 17448.934 | 200.972 | 203.27 | 1.204 | 1.165 |
| 55 | Burkina Faso | 16.9 | 971.299 | 972.754 | 17.421 | 17.465 | 1.794 | 1.795 |
| 56 | Ghana | 20.7 | 5109.591 | 5127.664 | 43.643 | 43.673 | 0.854 | 0.852 |
| 57 | Argentina | 31.9 | 217338.356 | 220929.146 | 2859.219 | 2866.866 | 1.316 | 1.298 |
| 58 | Colombia | 31.6 | 122317.762 | 123007.934 | 2740.851 | 2755.56 | 2.241 | 2.240 |
| 59 | Panama | 29.3 | 232223.876 | 237606.332 | 1943.255 | 1962.763 | 0.837 | 0.826 |
| 60 | Cuba | 41.4 | 99175.202 | 99454.451 | 760.779 | 760.779 | 0.767 | 0.765 |
| 61 | Ecuador | 28.3 | 58038.38 | 59392.36 | 1997 | 2002.277 | 3.441 | 3.371 |
| 62 | Venezuela | 28.3 | 19447.171 | 19528.686 | 206.03 | 206.913 | 1.059 | 1.060 |
| 63 | South Korea | 43.2 | 560818.7 | 667207.1 | 622.822 | 693.495 | 0.111 | 0.104 |
| 64 | Australia | 37.5 | 412017.8 | 442814.2 | 680.587 | 900.471 | 0.165 | 0.203 |
| 65 | New Zealand | 37.2 | 396786 | 458570.6 | 449.541 | 635.259 | 0.113 | 0.139 |
| 66 | Vietnam | 31.9 | 117379.7 | 118378.2 | 439.835 | 440.039 | 0.375 | 0.372 |
| 67 | Taiwan | 42.3 | 370284.7 | 428515.6 | 638.377 | 795.2 | 0.172 | 0.186 |
| 68 | Hong Kong | 45.6 | 350605 | 389150.1** | 1576.608 | 1895.5** | 0.450 | 0.487 |
| 69 | Bhutan | 29.4 | 79916.21 | 80128.37 | 26.839 | 26.839 | 0.0336 | 0.0335 |
| 70 | Tonga | 21.9 | 153574.1 | 157401.3 | 112.289 | 112.289 | 0.0731 | 0.0713 |
| 71 | Singapore | 42.8 | 390521.6 | 455261.1 | 303.529 | 332.09 | 0.0777 | 0.0729 |
| 72 | Montserrat | 43.7 | 317924.3 | 317924.3 | 1812.826 | 1812.826 | 0.570 | 0.570 |
| 73 | European Union | 44.4 | 397673.6 | 408494.6 | 2668.518 | 2763.03 | 0.671 | 0.676 |
| 74 | Europe | 42 | 325338.4 | 334708.4 | 2700.244 | 2788.427 | 0.830 | 0.833 |
| 75 | Africa | 18 | 9111.662 | 9201.963 | 181.186 | 181.545 | 1.989 | 1.973 |
| 76 | Asia | 31 | 58153.69 | 63664.54 | 325.065 | 345.988 | 0.559 | 0.543 |
| 77 | North America | 35 | 199059.8 | 207318.4 | 2580.998 | 2670.316 | 1.297 | 1.288 |
| 78 | South America | 31 | 152849.9 | 157558.3 | 3071.714 | 3104.744 | 2.010 | 1.971 |
| 79 | The world | 30.5 | 91468.7 | 96645.07 | 842.509 | 872.575 | 0.921 | 0.903 |

\* values calculated with the use of [22, 23]; \*\* values calculated with the use of [24, 25].
Zero-COVID countries are marked by green color. Aggregated characteristics are magenta.

## Notes

### Competing Interest Statement

The authors have declared no competing interest.

